# Effectiveness of Ghana’s COVID-19 policy responses and lessons learnt for the future: A multi-methods evaluation

**DOI:** 10.1101/2024.09.16.24313785

**Authors:** Shirley Crankson, Subhash Pokhrel, Nana Anokye

## Abstract

Ghana implemented various mitigating policies in response to the COVID-19 outbreak. This study examined the effectiveness of these policies to contribute to the ongoing discussions on proactive and pre-emptive interventions for similar future outbreaks.

A mix of qualitative and quantitative methods were used for the analysis. Data were drawn from multiple sources, including peer-reviewed and grey literature, and academic experts from Ghanaian universities. The data from the literature informed a questionnaire that was sent to independent academic experts to explore their opinions on whether the policies met their intended objectives. The experts’ opinions were collected on a 5-point Likert scale and from an open-ended question using an online data collection platform, Qualtrics. The data were evaluated using narrative synthesis, descriptive statistics and thematic analysis.

We identified and evaluated eight key COVID-19 policy responses in Ghana: (1) partial lockdown of epicentres; (2) COVID-19 public awareness campaigns; (3) ban on public gatherings; (4) COVID-19 vaccination; (5) border closures; (6) entry border COVID-19 screening; (7) incentives for healthcare workers (HCWs); and (8) the Ghana Alleviation and Revitalisation of Enterprises Support (GCARES). Two policies - the COVID-19 awareness campaigns and border closure - effectively improved public awareness of COVID-19 and helped to reduce COVID-19 case importation (median score ≥4).

Ghana’s COVID-19 public awareness campaigns and border closure policies could serve as a valuable model for informing proactive interventions to address future infectious disease outbreaks.

## Introduction

The COVID-19 outbreak has been one of the most consequential infectious disease outbreaks in the 21st century, affecting all facets of life, including global health systems, education and economies [1, 2]. However, while its horizontal experience has been comparable globally, its vertical impact has differed in many countries due to contextual boundaries emanating from multiple factors, such as socioeconomic and environmental variations [3]. Correspondingly, its mitigating policies have also been akin, to some extent, and differed in many regards globally to ensure their socioeconomic, cultural and environmental appropriateness and acceptability [4]. For example, many countries implemented a lockdown policy to curtail the spread of the outbreak; however, its operationalisation differed between countries owing to the earlier mentioned differences in contextual characteristics, including variations in housing structures, technological advancement, access to social amenities and economic activities [5]. A case in point is the lockdown intervention approach in Ghana and the UK, where the former implemented a partial lockdown and the latter a complete lockdown [6,7]. Apart from the similar policies with different operationalisation between countries, the contextual variations also accounted for very distinct and country-specific COVID-19 policies, like the Ghana Alleviation and Revitalisation of Enterprises Support (GCARES) policy, to ensure countries meet their specific pandemic and post-pandemic goals considering their COVID-19 experiences [8].

Notwithstanding the similarity and distinctiveness between countries regarding their COVID-19 policies, enhanced globalisation suggests that countries must continually evaluate their COVID-19 mitigating approaches to ultimately curb the outbreak on the global front [9]. One such evaluation could be reflecting and assessing the effectiveness of implemented policies to learn key lessons in order to inform policy guidelines for the continuous management of the outbreak or similar outbreaks in the future. The current COVID-19 literature is inundated with several such COVID-19 policies’ effectiveness analyses, with many of the analyses skewed towards many developed countries [10, 11]. Given the comparable policies globally, the findings from these analyses could offer helpful policy guides for other countries with scarcer literature representation. However, given the argued socioeconomic variations and country-specific COVID-19 policies, there is an urgent need for more comprehensive effectiveness analyses of country-specific COVID-19 policies, particularly from developing countries. These analyses can provide crucial evidence to support targeted and more tailored interventions in the event of another outbreak. They could also ensure a fair literature representation on COVID-19 mitigating strategies, which is fundamental for guiding the global mitigation agenda for similar outbreaks in the future. Therefore, this study, which evaluated the effectiveness of the COVID-19 policy responses in Ghana, a developing country with comparably scarce data in the literature on its COVID-19 policy assessments.

Ghana implemented several COVID-19 policies from 2020 to 2022 to lessen and avert any further dire repercussions of the outbreak [12]. These policies include a partial lockdown in the country’s COVID-19 disease hotspots, COVID-19 public awareness campaigns, enhanced testing capacities of earmarked COVID-19 testing centres and incentivisation of front-line workers [13]. Some of these policies have been evaluated in a few studies to ascertain their effectiveness and continuous relevance in the event of another outbreak. However, most of these studies primarily used a single approach, primarily qualitative approaches, to assess the policies’ effectiveness [12, 13, 14], offering nuanced evidence but limited robustness on the validity of the evidence. Further, most of the studies focused on the effectiveness of only a few of Ghana’s COVID policies [6, 15], offering insufficient data to enhance our understanding of the interconnectedness of Ghana’s multiple and simultaneously implemented COVID-19 policies and whether the policies’ interconnectedness impacted Ghana’s overall COVID-19 outcomes.

Therefore, this study was underpinned by a complementary multi-method approaches to diminish the subjectivity in the previous studies and enhance the validity of the evidence on the policies’ effectiveness [16]. As such, it combined a literature review approach with expert opinions (collected both qualitatively and quantitatively) to corroborate evidence on the effectiveness of Ghana’s COVID-19 policies. The literature review examined the existing evidence on the effects of Ghana’s COVID-19 policies and the quantitative and qualitative analyses leveraged independent expert perspectives on a 5-point Likert scale and open-ended questions to corroborate or challenge the literature review findings on the effectiveness of the policies. The study design required a framework for a standard or benchmark approach for demonstrating a policy’s’ effectiveness. Therefore, the policies’ objectives were used as benchmarks for assessing their effectiveness in the adopted multiple methods.

## Methods

This study was carried out in several stages as described below:

### Stage 1: Literature review

The study reviewed the grey and peer-reviewed literature to provide evidence on the effectiveness of Ghana’s COVID-19 policies in addressing the burden of the outbreak in Ghana. The data for the review was sourced from multiple sources: First, Scopus, the largest database of peer-reviewed journals and encompassing journals from other large databases [17], was searched for peer-reviewed articles evaluating Ghana’s COVID-19 policies using the keywords: ‘COVID-19’, ‘coronavirus’, ‘effectiveness’, ‘effect’, ‘impact’, ‘influence’, ‘policy’, ‘intervention’, ‘response’, ‘policy response’, ‘Ghana’. Second, the Google database was searched for generic articles using a combination of the above search terms. Third, specific websites, including the WHO, Worldometer and Government of Ghana (GoG) websites and local news agencies, were searched for reports on COVID-19 policy responses in Ghana. The GOG websites comprised Ghana Health Service (GHS), the Ministry of Health (MoH), the Ministry of Education (MoE), the Ministry of Information (MoI) and the Ministry of Finance (MoF) databases. The local media houses included Ghana Broadcasting Corporation (GBC), Citi FM, TV3 and Joy News, acknowledged and recognised local news agencies in Ghana that were involved in disseminating information on GoG’s responses to the COVID-19 outbreak. The multiple data sources contributed to enhancing the validity and reliability of identified information on Ghana’s COVID-19 policy responses. The search was first carried out from 30th March 2022 to 1st April 2022 and updated on 30^th^ July 2024. After the literature search, a narrative synthesis was done to compare information across multiple articles to provide evidence on a policy’s effectiveness. The whole review was guided by a logic model [18].

After the synthesis, a 3-point valuation measure was used to quantify a policy’s gains. The valuation, guided by the findings of the narrative synthesis, was necessary to provide an objective overview of the policy’s effectiveness. In the valuation, ‘0’ was given if literature review evidence showed that a policy did not effectively address any of its intended objectives, ‘1’ if at least one objective was met and ‘2’ when all objectives were met. Consequently, policies with a ‘0’ score were considered ineffective, those with ‘1’ were perceived as somewhat effective, and those with a score of ‘2’ were termed effective.

### Stage 2: Expert validation

Experts were invited to validate the effectiveness of the COVID-19 policies per their identified objectives in stage 1. Experts were defined in this study as academics, independent of the research team, with sufficient knowledge of the local context regarding Ghana’s COVID-19 policy landscape. The choice of academics as experts was informed by their documented attention to details, scientific rigour, and broader perspectives in developing, reviewing, and evaluating policies [19]. The academic experts were from universities in Ghana who were either teaching or researching in any health or economics disciplines. The two disciplines were informed by the nature of the COVID-19 policies in Ghana. A gatekeeper, who is affiliated to multiple universities in Ghana, was used to recruit the experts. The gatekeeper informed the experts about this study’s objectives and their expectations through a Participant Information Sheet (PIS) and furnished them with the link to the study’s questionnaire. The link and PIS were all shared through a broadcast study invitation email. The questionnaire was uploaded on Qualtrics, an online tool for data collection. Consent information was also embedded in the online questionnaire, and the experts could only record their responses after agreeing to the consent statement by clicking ‘agree’ on the online questionnaire. The link to the questionnaire was opened for the experts’ responses from 08/12/2022 to 16/01/2023. The questionnaire included sociodemographic questions and a question each on the objectives of the eight COVID-19 policies, as identified from the literature review. The experts rated the effectiveness of the policies on a scale of 1 to 5 (where 1 = not effective, 2 = not very effective, 3 = do not know, 4 = effective, 5 = very effective), except for the vaccination policy which was rated on a 1 to 3 scale (1 = no, 2 = maybe, 3 = yes) because it was still ongoing unlike the other policies, and the assessment aimed to examine its possible influence at the end of its implementation as per its prospective aim of inducing herd immunity. In addition to rating individual policies’ performance, the experts were given an open-ended field to record what they thought had contributed to a specific policy’s influence on a COVID-19 burden. Ethics approval for the expert’s recruitment and data collection was granted by the College of Health, Medicine and Life Sciences (CHMLS) Research Ethics Committee.

The questionnaire to collect expert opinions was a multi-item survey developed specifically for this study to allow items specific to the characteristics of Ghana’s COVID-19 policies, as no such questionnaire existed prior to this study. The questionnaire was informed by the findings of the literature review, suggestions from academics within the Department of Health Sciences, Brunel University London, and a review of the literature on standard approaches to questionnaire development to ensure content and face validity, respectively [20, 21]. A Cronbach’s alpha test was conducted to examine the reliability of the items on the survey questionnaire.

The analysis of the data on expert’s opinions involved several steps. Descriptive and thematic analyses were conducted to provide meanings to the experts’ responses. The descriptive analyses used frequencies, proportions, medians and Interquartile ranges (IQR) to summarise the experts’ characteristics. Policies with a median rating ≥4 were perceived as effective, and those with a median <4 were perceived as ineffective. An IQR value ≤1 was used to determine experts’ consensus on a policy’s effectiveness [22] The thematic analysis identified common themes from the participant’s responses to the open-ended question on what they think contributed to the outcomes of the COVID-19 policies. In the analysis, the responses from each expert were first read thoroughly to ensure familiarisation with the data. Codes were generated to represent phrases/sentences from the experts’ responses in the data familiarisation. The codes were then reviewed and observed for patterns regarding the policy’s effectiveness. Common codes from the experts were then combined into themes. The thematic analysis was conducted by SC and reviewed by NA to ensure that they accurately represented the experts’ responses. The final themes were defined in sentences to provide meanings to the experts’ responses and enhance understanding of the policies’ effect.

## Results

We evaluated eight key COVID-19 policies implemented in Ghana between 2020 and 2022. The policies were (1) partial lockdown of epicentres; (2) COVID-19 public awareness campaigns; (3) ban on public gatherings; (4) COVID-19 vaccination; (5) border closures; (6) entry border COVID-19 screening; (7) incentives for healthcare workers (HCWs); and (8) the Ghana Alleviation and Revitalisation of Enterprises Support (GCARES). The valuation measures, based on the literature review findings, found the public awareness campaigns, COVID-19 vaccination, border closures, entry border COVID-19 screening and the incentives for HCWs as ‘effective’, the partial lockdown and GCARES policies as ‘somewhat effective’ and the bans on public gatherings as ineffective. Table 1 summaries the literature review and valuation measures findings.

**Table 1:** Review findings and valuation score of Ghana’s COVID-19 policies’ effectiveness.

| **COVID-19 policy** | **Policy’s intended objectives** | **Review’s evidence on policy’s effect** | **Proportion (%) of studies reporting policy as effective** | **Valuation score** |
| --- | --- | --- | --- | --- |
| Partial lockdown of epicentres | Reduce spread of COVID-19 | Did not reduce COVID-19 spread [6; 14; 23; 24]<br>Unclear/inconclusive evidence [13] | 0/5 (0) | 1 (somewhat effective) |
|  | Enhance COVID-19 disease surveillance | Enhanced COVID-19 surveillance [24; 25; 26; 27] | 4/4 (100) |  |
|  | Scaling up COVID-19 | Scaled up COVID-19 testing and treatment | 3/5 (60) |  |

| **COVID-19 policy** | **Policy's intended objectives** | **Review's evidence on policy's effect** | **Proportion (%) of studies reporting policy as effective** | **Valuation score** |
| --- | --- | --- | --- | --- |
|  | treatment capacities | capacities [25; 27; 28]<br>Did not scale up COVID-19 testing and treatment capacity [26; 29]. |  |  |
| COVID-19 public awareness campaigns | Create awareness of COVID-19, its prevention and treatment protocols | Created awareness [13; 14; 25; 26; 30; 31; 32] | 7/7 (100) | 2 (effective) |
| Ban on public gatherings | Reduce COVID-19 transmission | Did not reduce COVID-19 transmission [ 23; 24; 27; 33] | 0/4 (0) | 0 (ineffective) |
| COVID-19 vaccination | Reduce risk and severity of | Reduced number of COVID-19 | 2/2 (100) | 2 (effective) |
|  | COVID-19 infections through vaccine-induced herd immunity | infections and deaths [23; 24] |  |  |
| Border closures | Reduce COVID-19 case importation | Reduced COVID-19 case importation by air [24; 34] | 2/2 (100) | 2 (effective) |
| Entry border COVID-19 screening | Detect and isolate COVID-19 cases | Detected and isolated over 7,000 COVID-19 active cases [24; 35] | 2/2 (100) | 2 (effective) |
| Incentives for healthcare workers | Widen workforce capacity | Increased in number of frontline healthcare workers [28; 36; 37] | 3/3 (100) | 2 (effective) |

| **COVID-19**<br>**policy** | **Policy's**<br>**intended**<br>**objectives** | **Review's evidence on**<br>**policy's effect** | **Proportion**<br>**(%) of**<br>**studies**<br>**reporting**<br>**policy as**<br>**effective** | **Valuation**<br>**score** |
| --- | --- | --- | --- | --- |
| GCARES | Stimulate<br>economic<br>recovery | • Stimulating economic recovery [38]<br>• Not simulating economic recovery [39; 40] | 1/3 (33) | 1 (somewhat effective) |

Thirty-four experts evaluated the eight COVID-19 policies, providing 272 main data points. Most of them were women (n = 25; 73.5%), aged 18 – 34 years (n = 17; 50%) and were from a health discipline (n = 30; 88.2%), including medicine, nursing and physiotherapy. They rated the public awareness campaigns, bans on public gathering, partial lockdown and border closures policies as effective (Median score ≥4), and the incentives for HCWs, COVID-19 entry border screening and GCARES policies as ineffective (Median score <4). There was consensus among the experts on the effectiveness of the public awareness campaigns (IQR =1). The reliability test showed a Cronbach’s alpha of 0.88, indicating a high internal consistency between the questionnaire’s items. Fig. 1 shows the median and IQR of the policies as rated by the experts. The vaccination policy is not included in Fig 1 as it was not rated on a 1 to 5 scale like the other policies. Also, only ten of the experts rated the effectiveness of the GCARES policy.

**Fig 1.**
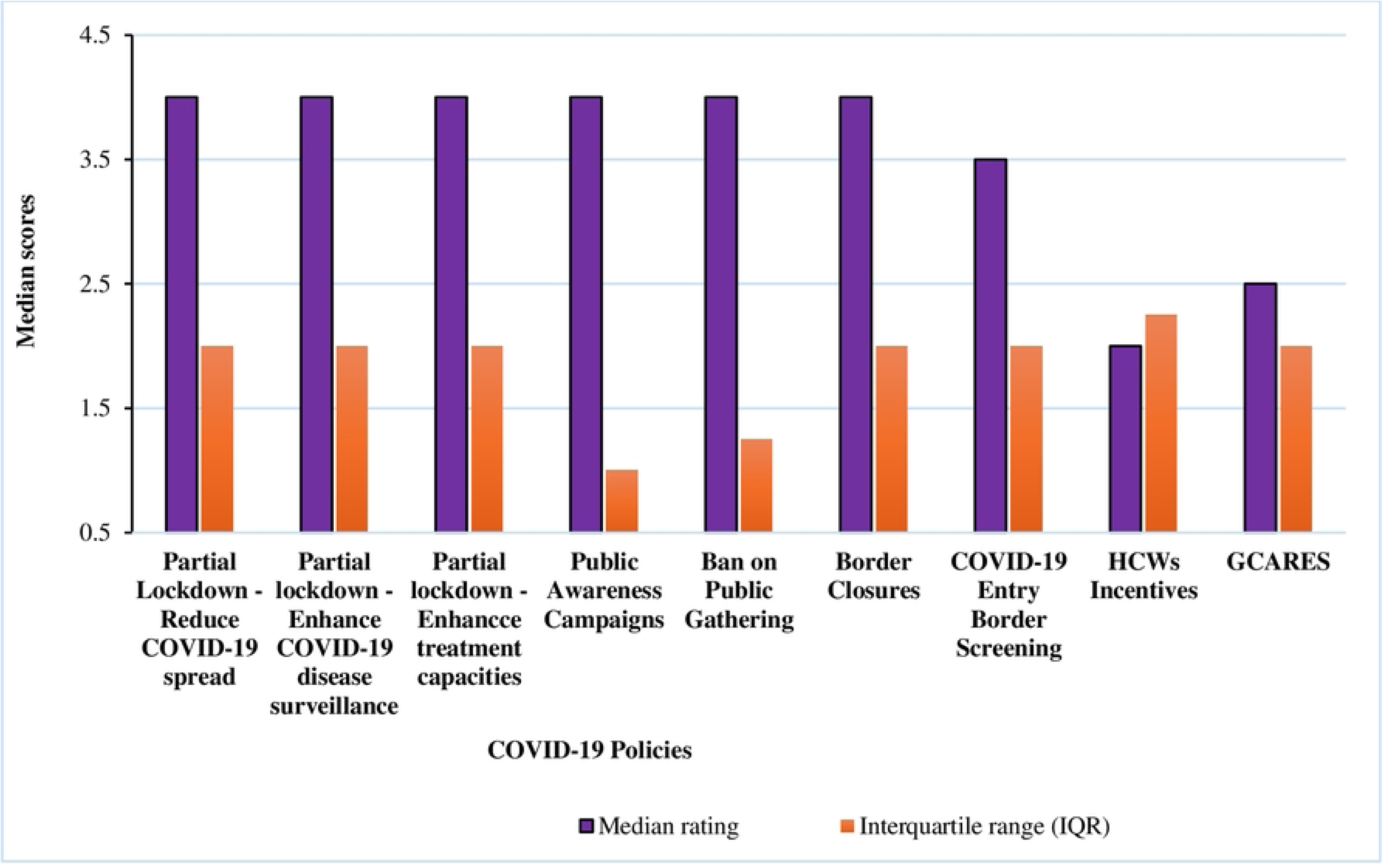
Experts’ ratings of the effectiveness of Ghana’s key COVID-19 policy responses.

The review findings for each policy and corresponding experts’ ratings of the policy’s effectiveness are presented below:

### (1) Partial lockdown of epicentres

A partial lockdown policy was implemented in two metropolises in Ghana – Greater Accra and Kumasi from 30th March 2020 to 20th April 2020 to reduce the burden of the COVID-19 disease [6, 41]. The policy allowed individuals in the targeted cities to access essential services, like food, pharmacy and banking services when needed, and members of the executive, judiciary, legislative, and media could operate [12, 13; 41]. The objective of the lockdown was to halt the spread of the virus (through movement restriction), enhance disease surveillance and scale up COVID-19 testing capacity [12, 42]. Before the policy, the number of COVID-19 cases was 152. This number increased to 1,042 on 20/04/2020 when the lockdown was lifted, indicating an 85% increase in the case count [23, 24]. The number of cases is reported to have increased steadily, even after the lockdown was lifted [23]. The increase in cases was attributed to testing backlog and intensified contact tracing, one of the objectives of the lockdown [28]. The contact tracing activity intensified COVID-19 disease surveillance [24], accounting for about 63% of active COVID-19 cases identified during the partial lockdown period [24, 25].

On the scaling up COVID-19 testing capacity objective, only two public laboratories, i.e., the Kumasi Centre for Collaborative Research (KCCR) and Noguchi Memorial Institute for Medical Research (NMIMR), were equipped to test for COVID-19 before and during the partial lockdown period [26]. The combined testing capacity of the laboratories before the lockdown was about 300 tests per day, and this doubled during the lockdown due to the adoption of a ‘pool testing system’, where tests were conducted in groups of 10s instead of individuals [34]. However, there were significant sample backlogs by the two labs during and post-lockdown, which affected the testing turnaround time [12, 28]. The pooling method was later abandoned when the case positivity rate increased [28]. Six hospitals were allocated for the management of COVID-19 during the lockdown, with one earmarked as the main treatment centre. Isolation and quarantine centres were also allocated in major cities during the lockdown period, with assistance from churches and private entities [28]. Capacity training was provided for the staff of these hospitals on COVID-19 testing, treatment, and appropriate use of Personal Protective Equipment (PPEs) [28]. These facilities, however, faced inadequate PPEs during the lockdown period, which affected their management of COVID-19 patients [26]. The partial lockdown was associated with unintended outcomes, such as job losses for individuals and institutions, social exclusion, and severe economic hardship for people with low incomes [14, 43]. The enhanced COVID-19 testing capacity of the country and some of the unintended outcomes informed the lifting of the partial lockdown in April 2020 [13, 42]. On the experts’ perceived effectiveness of the partial lockdown policy, twenty-two indicated that the policy was effective in enhancing COVID-19 disease surveillance (Effective: n = 17; Very Effective: n = 5), and nineteen of them said it was effective in reducing the spread of the virus (Effective: n = 15; Very effective: n = 4) and scaling up COVID-19 testing and treatment capacities (Effective: n = 15; Very effective: n = 4).

### (2) COVID-19 public awareness campaigns

COVID-19 public education and awareness campaigns were implemented in Ghana before the country recorded its first two COVID-19 cases on 12th March 2020 [42]. In the early stages of the outbreak, the campaigns were aimed to create public awareness of COVID-19 and ensure public adherence to COVID-19 prevention and management protocols [13]. As the disease progressed, the education campaigns were extended to include education on COVID-19 vaccination, with focus on demystifying the disease, ensuring social inclusion of recovered individuals and promoting COVID-19 vaccine uptake. The President was the key agent in the COVID-19 education campaigns. He encouraged the public to adhere to the COVID-19 preventive protocols and updated them on the government’s efforts to curtail the disease through frequent speeches through mass media [42]. There were several COVID-19 awareness campaigns in different local dialects on mass media across the country, and regular updates on the disease’s characteristics were made on government portals [14; 43]. The public could access COVID-19 information from multiple sources, including radio, TV and social media [31], with the internet being the major source of information [30]. The information included COVID-19 causes, symptoms, effects, and preventive measures [31]. Telecommunication companies also used push SMS to educate the public about COVID-19 [13]. Data showed that about 97% of Ghanaians knew about COVID-19 and the COVID-19 emergency centre [30], and the COVID-19 campaigns significantly influenced the awareness of the disease [32]. Notwithstanding, there were also reported cases of misinformation, mostly channelled through social media, friends and families [31, 44], and COVID-19-related stigmatisation [45].

The GoG and institutions issued directives to promote compulsory adherence to the COVID-19 preventive protocols, such as wearing nose masks and hand hygiene directives [46]. However, studies report low public adherence to the protocols, especially on hand washing, social distancing, and wearing face masks [46, 47, 48]. The low adherence to the protocols was linked to decreasing advocacy and awareness of COVID-19 by relevant agents, including the GOG [49]. There were campaigns on the benefits and safety of the COVID-19 vaccine [24; 26]. Some government officials took vaccines in public to create awareness of their safety [26]. Regular vaccine information was also provided on the GHS website. However, media campaigns on vaccine education were reportedly low [26]. Many experts (n = 30) rated the public awareness campaigns as effective (Effective: n = 21; Very effective: n = 9. The experts who rated it ineffective were divided equally between ‘Not effective’ (n = 2) and ‘Not very effective’ (n = 2).

### (3) Ban on public gatherings

The President of Ghana announced a COVID-19 informed public gatherings ban on 15th March 2020 to curtail the spread of the disease. The policy banned school activities, conferences, festivals, workshops, political rallies, religious activities, sporting events and all other social events for one month. However, private burials with a maximum of 25 attendees were permitted. The ban was initially imposed for four weeks and was extended until 5th June 2020, when the President eased the restrictions, citing low mortality and morbidity rates as reasons for the decision [42, 34]. Social activities, including conferences, workshops, and religious activities, were allowed with a maximum of a hundred persons following the lifting of the ban, and schools were reopened in batches for academic activities from 15th January 2021. However, in February 2021, the President re-introduced all the initial restrictions on public gatherings following a surge in COVID-19 cases [42]. The bans were finally lifted in March 2022.

During the public gatherings ban (from March 2020 to March 2022), the incidence of COVID-19 infections fluctuated. However, the highest number of daily new cases (n = 2,521) since the outbreak began was recorded within the ban period [23, 24]. It is argued that the reported high daily COVID-19 incidences during the ban period were possibly influenced by spillovers from before the ban’s imposition due to the observed delays in laboratory testing and case reporting [12, 50]. Evidence suggests that COVID-19 transmission did not decline following the bans on public gatherings, and relaxing the ban did not increase COVID-19 transmission [33]. Reopening schools, however, increased the COVID-19 transmission rate in some regions of Ghana [33]. Several reports also allude to ban violations, especially by religious groups and government officials during the ban period, which may have increased COVID-19 transmission [13, 14, 45, 51]. Most experts (n = 24) rated the ban on public gatherings policy as effective (Effective: n = 17; Very effective: n = 7) in reducing the spread of the virus. Of those that said the policy was ineffective, six rated it as ‘not very effective’, and two rated it as ‘not effective’.

### (4) COVID-19 vaccination

Ghana implemented a mass COVID-19 vaccination intervention on 1st March 2021 after receiving 600,000 doses of AstraZeneca on 24th February 2021. The intervention was to reduce the risk and severity of COVID-19 infections through vaccination-induced herd immunity [24, 52]. It targeted a 60% full vaccination (2 doses of a vaccine) of Ghanaians aged ≥15 years by the end of June 2022 to achieve herd immunity [42]. The proportion of the targeted Ghanaians vaccinated as of 9th September 2024 is 56.7% [24], 3.3% shy of the June 2022 target. The total number of COVID-19-related deaths as of 1st March 2021, when the mass vaccination began, was 607. This number had increased to 1,462 as of 7th April 2024 (The last COVID-19 death updates by GHS) [24, 53], representing 855 deaths. The 607 reported deaths were in the space of 12 months (between 12th March 2020 and 1st March 2021), representing approximately 51 deaths per month. The 855 additional deaths were also in the space of 37 months (2nd March 2021 to 7th April 2024), representing about 23 deaths per month. This data suggests that the number of monthly COVID-19-related deaths reduced by about 55% since the vaccination policy began. On the experts’ perspectives of whether the vaccination policy could achieve vaccine-induced herd immunity, nineteen (56%) said the vaccine policy ‘may’ reduce the COVID-19 burden through herd immunity, two (6%) said the vaccine would not achieve this objective, and thirteen (38%) of them were certain that the policy would achieve the objective.

### (5) Border closures

Ghana announced a non-admittance restriction on other nationals travelling from countries with over 200 confirmed COVID-19 cases, except those with resident permits, on 15th March 2020 to reduce COVID-19 case importation [54]. This restriction continued until 22nd March 2020, when all borders, including air, sea and land, were completely closed to all outbound and inbound travellers; however, the movement of Cargo, essential goods and supplies was not restricted [55]. The first two COVID-19 cases in Ghana were imported cases, which increased to 105 by mid-March 2020, necessitating the border closure intervention [23, 24]. The borders were opened for air travel on 1st September 2020, but the restrictions on land borders remained until April 2022. Data shows that 705 imported cases were recorded from March 2020 to December 2020 [28], suggesting that about 600 COVID-19 cases were imported after lifting the air travel restrictions, i.e., from September to December 2020, as 105 imported cases had already been reported pre-border closure. By estimation, about 150 cases were imported monthly for the first four months after lifting the border closure restrictions. This estimation could also suggest that about 150 cases per month, representing about 750 cases for the period of the ban imposition (March – September; 5 months), were prevented [24, 34]. Most of the experts (n = 20) said the border closure policy effectively prevented COVID-19 case importation (Effective: n = 13; Very effective: n = 7). Three did not know whether the policy prevented case importation, and eleven rated the policy as ineffective per its targeted objective.

### (6) Entry border COVID-19 screening

Prior to the re-opening of air borders, Ghana introduced compulsory COVID-19 screening at its main international airport, the Kotoko International Airport (KIA), in September 2020 [42]. The policy aimed to detect and isolate imported COVID-19 cases at entry points to prevent a case-importation-induced increase in COVID-19 prevalence. In addition to the compulsory airport testing, passengers were mandated to present a negative PCR test result from their country of embarkment [28]. The compulsory entry border screening was suspended when the land borders were opened for outbound and inbound travel in March 2022 [42]. Infographics from GHS showed that the COVID-19 testing at KIA identified and isolated over 7,000 active cases during the policy’s implementation period [24]. The experts’ ratings of the border screening policy’s effectiveness were spread equally between effective (Total = 17; Effective: n = 15; Very effective: n = 2) and ineffective/unsure of effectiveness (Total = 17; Not effective: n = 2; Not very effective: n = 10; Don’t know: n= 5). But when fragmented, most experts (n = 15) rated the policy as ‘effective’ rather than ‘not very effective’ (n = 10).

### (7) Incentives for HCWs

Ghana announced incentive packages for healthcare workers in March 2020 to widen its workforce to fight the COVID-19 outbreak [37]. The incentives included about US$60,000 in insurance coverage per person, free transportation, 50% of the basic salary allowance for all frontline workers, and tax-free salaries on employee emoluments for all health workers [37]. These financial packages were rolled out from April 2020 to December 2020 [56, 57]. In addition, over 45,000 healthcare workers were recruited from March 2020 to November 2022, increasing the health worker capacity by about 35% [36]. By the end of 2020, Ghana had spent about US$35 million on health workers’ financial packages and recruitment to sustain and boost its workforce capability against the COVID-19 outbreak [36]. The experts who said the HCWs incentives policy was effective (Total = 14; Effective: n= 11; Very effective: n = 3) were fewer than those who said the policy was ineffective (Total = 18; Not effective: n= 8; Not very effective: n = 10) in widening the human resource capital against COVID-19.

### (8) GCARES

Ghana implemented a GCARES policy in May 2020 to stimulate economic recovery from the COVID-19 impact [58]. The three- and half-year program was rolled out in two phases [58]. The first phase focused on revamping the economy through tax exemptions, reduced cost of essential services and provision of loans up to 600 million cedis with two years repayment schedule for informal and formal Micro, Small and Medium-scale Enterprises (MSMEs) [58]. As of 21st May 2020, about 8,000 applicants had registered to access the loan to revamp their businesses [59]. The first phase ended in July 2020. The second phase, launched for three years (2021-2023), aimed to transform Ghana’s economy through revived industries, such as manufacturing, construction, digitalisation and agri-business [58]. Data shows that the Gross Domestic Product (GDP) in the fourth quarter of 2021 (7%) was higher than 2020’s GDP (4.3%), indicating marginal economic growth, which could have been influenced by the GCARES policy [38]. Of the ten experts who rated the effectiveness of the GCARES policy, five said the policy was ineffective (Not effective: n = 3; Not very effective: n = 2) in stimulating economic recovery from the COVID-19 impact. Four of the remaining five did not know the policy’s effectiveness, and one said it was effective.

On the open-ended responses on the rationale for the policies’ effectiveness rating, ten of the experts gave reasons for their effectiveness ratings of the COVID-19 policies. Their reasons are summarised in Table 2 below. Many (n = 4) of them commented on the ban on public gatherings policy and their reasons included:

**Table 2:**
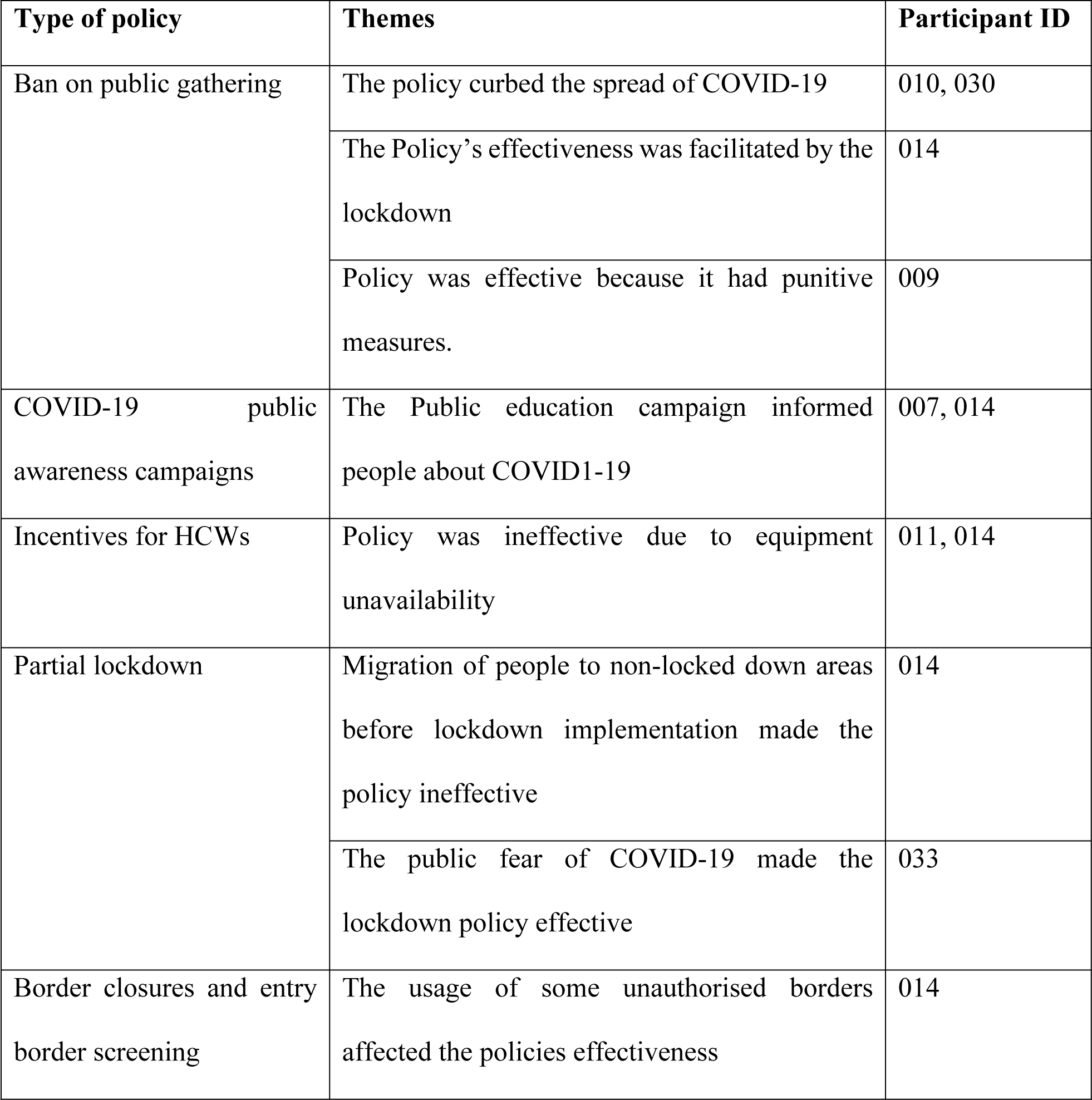
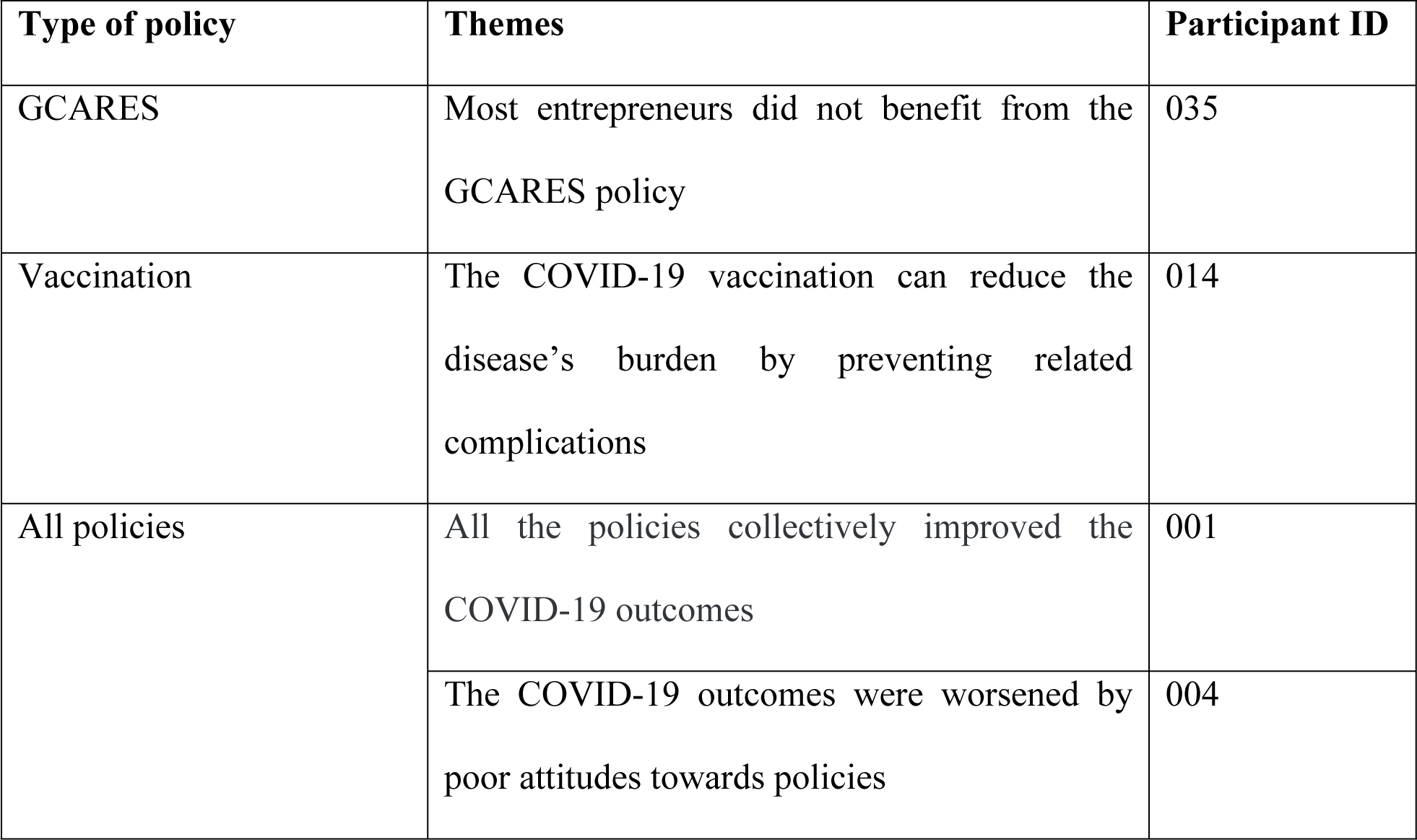
Thematic findings of the experts’ reasons for their policies’ effectiveness ratings.

*“On the ban of the public gathering, it really helped to curb the spread of the virus. It was actually one of the effective measures employed” (Participant 010)*.

*“The restriction on public gathering was very effective. This was because churches and club houses were closed, and anyone seen disobeying were punished” (Participant 009)*.

On other policies, one explained that the *“the closure of the borders in a way helped to reduce spread of Covid-19, but it affected traders a lot and people used unauthorized borders to go about their business. So, at the end it wasn’t effectively controlled and not everyone got tested” (Participant 014)*.

## Discussion

To the best of our knowledge, this is the first study to examine the effectiveness of key COVID-19 policies in Ghana, by first using a robust scrutiny of the literature and then validating the findings in a rigorously structured expert poll. The literature review assessment of effective (public awareness campaigns, COVID-19 vaccination, border closures, entry border COVID-19 screening and HCWs), somewhat effective (GCARES and partial lockdown) and ineffective (bans on public gatherings) policies was consistent with the experts’ opinion for some policies and inconsistent with others. For example, agreement was obtained between the review findings and experts’ ratings regarding the COVID-19 public awareness campaigns as being effective but there was disagreement on the effectiveness of the partial lockdown, ban on public gatherings, HCWs incentives, GCARES and entry border COVID-19 screening policies. The disagreement could emanate from the policies’ characteristics and operationalisation, which blurs objective evaluations. For example, some of the policies were complementary, had similar intended outcomes and were implemented around the same time, therefore obscuring possible attribution of policy outcomes to specific policies. A case in point is the partial lockdown and bans on gatherings policies which were implemented around the same time and had the same intended outputs of reducing the spread of COVID-19. This similarity restricted estimating actual outcomes attributable to either of the policies, limiting precise objective assessments. This limitation could have contributed to the variations in the results between experts’ opinions and the initial ratings. Arguably, the study could have addressed this attribution limitation with a period analysis of their outcomes [60]. However, given that these policies were implemented concurrently, there were insufficient data for such an analysis.

Unlike the other policies, the vaccination policy was assessed prospectively by the experts, given that it is still in force. Therefore, its experts’ findings may not represent the policy’s present gains. Nonetheless, its literature review findings demonstrate a reduction in the number of COVID-19 infections and related deaths since the vaccination intervention started. This reduction is not specific to any vaccine type and no empirical data was found to attribute this reduction to the vaccination intervention. However, a recent modelling study showed that Ghana’s vaccination intervention could reduce symptomatic COVID-19 infections in adults aged 25 to 64 years by about 7% [61]. This evidence suggests that Ghana could enhance its vaccination uptake to continue reducing its COVID-19 infections and deaths.

The literature review finding on whether the vaccination policy achieved herd immunity was comparable to the experts’ perspectives. Ghana did not achieve its herd immunity objective in June 2022, even almost two years after, potentially due to vaccine unavailability and hesitancy [52]. Studies have reported vaccine hesitancy in Ghana and have attributed it to poor knowledge, anxiety, conspiracy theories, safety concerns and misconceptions [62–64]. This observation, however, is not isolated to Ghana, as several studies from other jurisdictions have also reported COVID-19 vaccine hesitancy [65–67]. When positioned within behavioural models, it could be explained that vaccine uptake among populations could be influenced by a multidirectional interplay of complex sociocultural, religious, and behavioural factors [68], warranting a comprehensive intervention to prevent hesitancy. Policymakers in Ghana could, therefore, explore these factors to scale-up its vaccination intervention uptake and address any vaccine hesitancy that could impede efforts at meeting its herd immunity target.

The review showed that the entry border COVID-19 screening policy helped isolate and quarantine travellers who tested positive for COVID-19 on arrival [35]. Consequently, those testing negative for the virus were allowed entry into the country [35]. Therefore, it could not be established whether the policy was sufficient in curtailing COVID-19 transmission from imported cases, particularly as a negative test at the point of entry may not indicate a ‘true’ negative COVID-19 status, given the influence of viral incubation periods on test results [69], and it was unclear in the literature whether the travellers with negative test results on arrival were required to have a repeated test within specified time to confirm the arrival test outcome.

The partial lockdown policy did not reduce COVID-19 spread per the literature review findings, and though the experts rated it effective in reducing COVID-19 transmission, they did not achieve consensus. The increased COVID-19 cases during the lockdown [23] could have been from the not-lockdown areas. However, data from GHS [24] shows reported COVID-19 cases from the lockdown areas, suggesting viral spread in the epicentres during the policy period. The viral spread could have been facilitated by hindered early identification and isolation of cases due to the delayed testing turnaround time experienced during the lockdown because of the lower testing capacities in Ghana [28, 70]. It could have also resulted from the reported scaling down of contact tracers at some point during the lockdown imposition [25]. Like the partial lockdown, there was no consensus among the experts on the effectiveness of the bans on public gatherings policy, and the review found it ineffective. Given the Ghanaian socioeconomic context, it was probably impossible to curtail COVID-19 transmission through the imposition of a public gathering ban. This is because it is impractical to attain complete adherence to such a ban at all population levels due to socioeconomic inequalities [13]. Like many developing countries, individuals at the bottom of the economic pyramid in Ghana face myriad challenges, including low purchasing power, shared and poor sanitation facilities and housing conditions [71]. These challenges may limit their tendency to avoid public gatherings and practise social distancing [72].

There are several lessons learnt from this study for handling future pandemics of the nature and scale of COVID-19. First, relevant policymakers in Ghana could consider the country’s socioeconomic and cultural fabrics, such as education, traditions, religion and housing characteristics, when implementing policies like lockdowns and bans on public gatherings to ensure their optimal impacts in the event of another pandemic. When needed to flatten the transmission curve, Ghana could consider a lockdown intervention when the number of infections exceeds treatment capacities [73]. However, this decision must be complemented with scaled-up testing capacities and well-equipped treatment facilities to avoid prolonged lockdown periods, which could have dire economic consequences [25]. Second, given Ghana’s economic situation, which usually affects its provision of equipment and resources for health service delivery, a public-private partnership could be helpful to ensure an uninterrupted and adequate supply of resources to manage pandemics. The government could also support hospital facilities to generate internal funds through grants, such as clinical grants, to ensure their self-reliance, which could boost their preparedness for emergencies like pandemics.

Third, Ghana’s government could equip and support relevant state agencies and appropriate private organisations to improve its policy regulations and enforcements strategies. This support is necessary as the evidence suggests that enhanced policy enforcement strategies are crucial to mitigate the burden of any similar future outbreak in Ghana [13]. Fourth, Ghana could adopt its COVID-19 awareness campaigns as a model communication and awareness strategy for educating the public in future infectious disease outbreak management, as evidence supports its effectiveness for pandemic management [74]. The policy could be strengthened further by blocking sources of misinformation. Finally, global systems could enhance their efforts to promote COVID-19 herd immunity by ensuring equitable distribution of COVID-19 vaccines for the benefits of developing countries like Ghana. They could also consider strategic partnerships with social media platforms to control spread of misinformation during disease outbreaks.

Our study used data triangulation to advance the knowledge of the effectiveness of Ghana’s critical COVID-19 policy responses. Leveraging the advantages of this approach allowed our study to contribute more reliable evidence of the policies’ effectiveness. In addition, we integrated quantitative and qualitative approaches, offering complementary approaches that could diminish each approach’s weaknesses while maintaining their strengths. However, as associated with qualitative analysis, this study is possibly limited by some subjective data interpretations, which could have implications for interpreting our findings. However, given the data triangulation herein, this limitation could be subdued. Our study was also faced with attribution of effects limitations, given the similar characteristics of the policies and the non-availability of comparable data to address counterfactuals. For example, the researchers could not attribute the identified COVID-19 outcomes to any single policy, as most of the evaluated policies in this study were implemented concurrently. Therefore, by inferences, the COVID-19 outcomes discussed here could be interpreted as the outcome of all the policies combined. Also, the relatively small number of experts in this study limits the generalisation of the findings as a reflection of the viewpoints of all academic experts in Ghana. Further, the evaluations focused mainly on the policies’ direct effects, limiting comprehensively accounting for their indirect effects.

Comparing our findings to the literature, no study was found to have evaluated all the policies herein at a go, as done in this study. Therefore, the findings presented here could not be compared to the literature as a composite. However, when defragmented, the findings on the partial lockdown policy and the public awareness policy are consistent with similar studies [6; 32].

## Conclusions

Ghana’s COVID-19-related public awareness campaigns and border closure policies effectively informed the public about COVID-19 and contained COVID-19 case importation, respectively. All the identified eight COVID-19 policies contributed to the COVID-19 outcomes in Ghana. Future studies could expand on the current understanding by exploring more robust data and approaches to examine the effectiveness of Ghana’s COVID-19 policies.

## Data Availability

The data accessed from the experts and analysed in this study is publicly available at Figshare and is available from: https://doi.org/10.6084/m9.figshare.22640203.v1 All data included in the literature review are contained in the manuscript

## Acknowledgments

The authors wish to thank the Ghana Scholarships Secretariat for their immense support towards this research, and all the experts who participated in this research.

